# Pediatric Hypertension Gaps: Longitudinal Analysis and Social Drivers of Health

**DOI:** 10.64898/2026.06.30.26356982

**Authors:** Abbas H. Zaidi, Nathanael Rehmeyer, Erica Sood, Sarah De Ferranti, Gumpili Sai Prashanthi, Benjamin Brewer, Katherine Campbell, Joanne Lopes, Colleen Witherell, Jonathan Miller, Anne Kazak

## Abstract

**Background:** Pediatric hypertension (HTN) remains underdiagnosed despite established guidelines. Prior studies evaluating social drivers of health (SDOH) relied on diagnostic codes or single-visit blood pressure (BP) measurements, limiting identification of persistent BP elevation. We evaluated longitudinal BP patterns and associated SDOH among children with continuity of care.

**Methods:** We conducted a retrospective cohort study of children aged 6–17 years with ≥3 primary care visits between 2017 and 2024 within a large healthcare network. BP was classified according to guidelines. Multivariable logistic regression evaluated associations between persistent abnormal BP (≥3 abnormal readings, would meet guideline-based diagnosis for HTN) and stage 1/2 HTN with demographic, clinical, and neighborhood-level factors, including Area Deprivation Index (ADI), Child Opportunity Index (COI), and insurance instability.

**Results:** Among 71,683 children, 2,911 (4.2%) had persistent abnormal BP, whereas only 848 (1.2%) had a documented HTN diagnosis. Obesity, age ≥13 years, male sex, prematurity, and insurance instability were associated with abnormal BP and stage 1/2 HTN. Higher ADI quartiles were associated with increased odds of abnormal BP (Q3: OR 2.48) and stage 1/2 HTN (Q3: OR 4.89). Higher COI socioeconomic opportunity was associated with lower odds of abnormal BP, whereas higher educational opportunity was associated with higher odds; these associations were not observed for stage 1/2 HTN.

**Conclusions:** Among children with continuity of care, substantial gaps in HTN recognition persist despite repeated opportunities for diagnosis. SDOH factors remained associated with BP abnormalities, supporting the need for system-level and community-based strategies to improve HTN detection.

## INTRODUCTION

Pediatric hypertension (HTN) remains underdiagnosed despite established screening guidelines, with fewer than 25% of children who meet American Academy of Pediatrics (AAP) criteria receiving a formal diagnosis.^1–14^ As a result, reliance on diagnostic codes likely underestimates the true burden of disease and fails to capture children with persistently elevated blood pressure (BP) who remain unrecognized in clinical practice.^11,12,15^

Children with a documented HTN diagnosis have been shown to differ by social drivers of health (SDOH) and neighborhood context; higher HTN rates observed among those living in more socioeconomically deprived areas and among those with public insurance.^16–21^ Similarly, cross-sectional studies have demonstrated sociodemographic differences in elevated BP.^21,22^ However, these approaches rely on diagnostic codes or single-visit measurements and cannot distinguish transient elevations from persistently elevated BP at multiple visits, which meets clinical criteria for HTN diagnosis based on guidelines.^15,18,21–24^ Less is known about children who meet criteria for HTN based on repeated BP measurements but remain undiagnosed.

Neighborhood measures such as the Area Deprivation Index (ADI) and Child Opportunity Index (COI) capture complementary dimensions of structural context.^21,22,25–29^ However, their relationship to longitudinal BP patterns and HTN recognition among children who maintain continuity of care has not been well characterized.

To address this gap, this study focused on children with established continuity of care and sufficient BP measurements to assess HTN using guideline-based diagnostic criteria, thereby minimizing the effect of access-related barriers. We aimed to (1) determine the prevalence of HTN based on longitudinal BP patterns and (2) evaluate associations with SDOH, including insurance instability, ADI, and COI. We sought to identify system-level gaps in HTN detection among underdiagnosed HTN patients with continuity of care. In this study, race and ethnicity were considered social constructs that may reflect differential exposure to structural and SDOH rather than inherent biological differences. In this study, we assess the effects of neighborhood SDOH, beyond access to care, among patients with elevated BP meeting clinical criteria for HTN.

## METHODS

### Study Design and Data Source

This retrospective cohort study utilized electronic medical record (EMR) data from a large tertiary care hospital network with primary care clinics located in Delaware, Pennsylvania, Maryland, and New Jersey. The study period extended from August 2017 through November 2024, corresponding with the publishing of the 2017 American Academy of Pediatrics (AAP) HTN guidelines.^2,7^

Institutional Review Board approval was obtained prior to data extraction. Informed consent was waived due to the retrospective nature of the study and use of de-identified data. Data, analytic methods, and study materials are available from the corresponding author upon request.

### Study Population

Eligible participants included pediatric patients aged 6 to 17 years who were seen for well-child primary care visits during the study period and had at least three primary care clinic visits during the study period. Patients with less than three visits were excluded as they were likely not continuity patients within the healthcare system and did not have three visits to make a diagnosis of HTN. This approach was intended to reduce misclassification from transient BP elevations and approximate guideline-based diagnostic criteria requiring persistent BP elevation for at least three visits. The lower age cutoff of 6 years was selected due to recognized variability in BP measurement accuracy in younger children and the increased likelihood of secondary HTN in early childhood.^2,7^ Patients were excluded if they had documented diagnoses associated with developmental delay or autism spectrum disorder, given the potential for falsely elevated BP readings related to behavioral distress during clinical encounters.^30,31^ Patients younger than 13 years without recorded height data were excluded because BP percentiles could not be calculated. Additionally, patients residing outside the four study states were excluded as these patients likely did not represent primary care continuity patients and did not have adequate longitudinal data for analysis.

### Hypertension Diagnosis and Blood Pressure Classification

To assess longitudinal BP trends, only the first three eligible visits per patient during the study period were included. Patients who demonstrated mixed BP patterns without meeting predefined classification criteria (described below) were excluded to ensure consistent longitudinal HTN diagnosis categorization.

BP categories were assigned at each visit exactly in accordance with the 2017 AAP HTN guidelines.^2,7^ For children younger than 13 years, BP classification was based on age-, sex-, and height-specific percentiles derived from standardized AAP reference tables.^2,7^ In this age group, normal BP was defined as <90th percentile, elevated BP as 90th to <95th percentile, and hypertensive BP as ≥95th percentile. For children aged 13 years and older, BP classification was based on absolute thresholds: normal BP was defined as systolic BP (SBP) <120 mmHg and diastolic BP (DBP) <80 mmHg; elevated BP as SBP 120–129 mmHg and DBP <80 mmHg; stage 1 HTN as SBP 130–139 mmHg or DBP 80–89 mmHg; and stage 2 HTN as SBP ≥140 mmHg or DBP ≥90 mmHg as outlined in the guidelines.^2,7^ If height was missing at a given visit for children under 13 years, the most recent recorded height within the preceding 180 days was used to calculate BP percentiles. For visits in which multiple BP measurements were recorded on the same day, values were aggregated to represent a single BP measurement in accordance with guideline-recommended clinical practice.

### Patient-Level Blood Pressure Classification

Patients were categorized into mutually exclusive groups as follows:

- **Normal BP group:** All available visits (≥3 visits) were classified as normal.
- **Abnormal BP group:** Three consecutive separate visits with BP values in the elevated, stage 1 or stage 2 BP ranges, with the recent visits continuing to show abnormal BP.
- **Stage 1 or Stage 2 HTN group:** Three consecutive separate visits specifically meeting stage 1 and/or stage 2 HTN criteria, with the recent visits continuing to show abnormal BP.

Patients with only one or two abnormal readings without a third confirming abnormal BP visit were excluded as they did not meet criteria for continuity of care. Similarly, patients demonstrating mixed BP patterns (i.e., a combination of visits with normal and abnormal BP without three consecutive abnormal readings) were excluded from analysis to limit patients with potentially erroneous elevated BP or white coat HTN.

### Hypertension Diagnostic Code

The presence of a HTN diagnosis was determined using the International Classification of Diseases, Tenth Revision (ICD-10) code I10 (Primary Hypertension) and associated codes documented in the EMR. Diagnostic codes were not used to assign BP classification groups outlined above. Instead, the presence of an ICD-10 HTN diagnosis was analyzed as a separate variable to evaluate potential under recognition of HTN among children who met BP-based criteria.

### Clinical Variables

Age was extracted based on the clinic visit at the time of BP measurement and was used to calculate BP percentiles for that visit. Age at baseline was further dichotomized as <13 years and ≥13 years to align with the transition from percentile-based to absolute BP thresholds per guidelines.^2,7^ Age was then used for analysis based on the most recent clinic visit. Body mass index (BMI) percentiles were calculated using Centers for Disease Control and Prevention growth charts. BMI categories were defined as underweight (<5th percentile), healthy weight (5th to <85th percentile), overweight (85th to <95th percentile), and obese (≥95th percentile). If BMI category varied across visits, the most frequently observed category was assigned. Prematurity was defined as birth prior to 37 weeks gestation.

### Demographic and Social Drivers of Health Variables

Race and ethnicity were self-reported at clinical visits. Original EMR race categories were combined with ethnicity into four mutually exclusive groups: White non-Hispanic, White Hispanic, non-White non-Hispanic, and non-White Hispanic. Sex was categorized as male or female. English proficiency was defined based on primary language documentation and categorized as English versus non-English. Insurance was categorized as public (Medicaid, Managed Medicaid, Medicare, or Tricare) or private (all other payors). Insurance changes (as a marker for insurance/financial instability) were noted as having two or more changes of a unique payor within 1 year of recorded medical history. If multiple home zip codes were documented during the study period, the zip code from the most recent visit was used for neighborhood-level linking. Household caregiver structure was assessed using the number of caregivers documented in the electronic health record and categorized as one caregiver or ≥ 2 caregivers.

### Child Opportunity Index

Neighborhood-level opportunity was assessed using the COI 3.0, developed by the Institute for Children, Youth, and Family Policy at Brandeis University.^27^ COI is a multidimensional composite measure designed to capture neighborhood conditions that influence child health and development. It incorporates 29 indicators drawn from publicly available data sources, including the U.S. Census Bureau, National Center for Education Statistics, U.S. Department of Agriculture, and Environmental Protection Agency. These indicators reflect structural and contextual neighborhood characteristics such as household income, employment rates, health insurance coverage, school quality, environmental exposures, and access to community resources.^27–29^

COI 3.0 organizes these indicators into three domains: education, health and environment, and socioeconomic. Composite scores are standardized and range from 1 to 100, with higher scores representing greater neighborhood opportunity. Scores are normed at metropolitan, state, and national levels.

Patients were assigned COI values based on their residential zip code at the most recent clinical encounter during the study period. Zip code linking was performed using the *zipcodeR* package in R (R Foundation for Statistical Computing, Vienna, Austria). Children whose recorded home zip codes were located outside Delaware, Pennsylvania, Maryland, or New Jersey were excluded from COI-based analyses to maintain geographic consistency within the study region.

Overall COI scores were categorized into three groups to improve interpretability and model stability: low/very low (<40), moderate (40 to <60), and high/very high (≥60). Domain-specific COI scores for education, health and environment, and socioeconomic also were included in multivariable regression models to evaluate whether distinct dimensions of neighborhood opportunity were independently associated with BP outcomes.

### Area Deprivation Index

Neighborhood socioeconomic disadvantage was additionally assessed using the 2023 ADI, obtained from the Neighborhood Atlas.^26^ The ADI is a validated composite measure developed using American Community Survey (ACS) 5-year data and supported by the Health Resources and Services Administration (HRSA). It ranks neighborhoods at the Census Block Group (CBG) level based on indicators of income, education, employment, and housing quality. Higher ADI percentiles reflect greater neighborhood deprivation.

Because ADI is reported at the CBG level and patient residence was available at the zip code level, a multistep linking process was performed. First, CBG-level ADI rankings were aggregated to the census tract level. Tract-level ADI values were then merged with the U.S. Department of Housing and Urban Development (HUD) ZIP–TRACT crosswalk file to associate each tract with corresponding zip codes. Finally, ADI values were calculated at the zip code level by averaging tract-level ADI rankings for all tracts linked to a given zip code. Each patient was assigned a national ADI percentile ranking based on their most recent recorded residential zip code during the study period, consistent with prior methodology.^32^ For statistical analyses, ADI was treated as an ordinal/categorical variable representing national percentile rank and divided into quartiles based on the total population studied.

### Statistical Analysis

All statistical analyses were performed using R version 4.3.1. Summary statistics were calculated overall and stratified by BP classification. Continuous variables were summarized using means and standard deviations, and categorical variables using frequencies and percentages. Group comparisons were conducted using chi-square tests for categorical variables and t-tests for continuous variables.

Two multivariable logistic regression models were constructed to evaluate associations between clinical and SDOH variables and BP outcomes: (1) abnormal BP (three consecutive elevated or hypertensive readings) versus normal BP, and (2) stage 1/stage 2 HTN (three consecutive readings with BP values meeting criteria for stage 1 or stage 2 HTN) versus normal BP. All models adjusted for age group, sex, race and ethnicity, English proficiency, insurance type, insurance changes, BMI category, prematurity, national ADI, and COI domain categories. Adjusted odds ratios (OR) with 95% confidence intervals (CI) were reported. Model discrimination was assessed using C-statistic. A two-sided α level of 0.05 was considered statistically significant.

## RESULTS

### Patient Characteristics

Based on study timeframe (2017-2024), a total of 118,673 children were in the initial cohort, of which 3,084 (2.6%) had a documented diagnosis of HTN based on ICD codes. After restricting the cohort to children with at least three consecutive visits based on study criteria (71,683), only 854 (1.2%) had a diagnosis code for HTN, while 2,911 (4.2%) met diagnostic criteria for HTN based on guidelines (Table 1). Among children with abnormal BP based on study definition, 671 (23.1%) had a coded HTN diagnosis.

**Table 1.** Social demographics and clinical parameters for participants based on BP.

|  | **Total n (%)** | **Normal BP n (%)** | **Abnormal BP n (%)** | **p-value** |
| --- | --- | --- | --- | --- |
|  | n = 71683 | n = 68772<br>(95.8) | n = 2911<br>(4.2) |  |
| **Social Demographics** |  |  |  |  |
| **Male Sex** | 33217 (46.3) | 31377 (45.6) | 1840 (63.2) | <0.001 |
| **Adolescent Age ($\geq 13$ years)** | 31510 (44.0) | 28871 (42.0) | 2639 (90.7) | <0.001 |
| **English Proficiency** | 67125 (93.6) | 64480 (93.8) | 2645 (90.9) | <0.001 |
| **$\geq 2$ Caregivers** | 67271 (93.9) | 64490 (93.8) | 2781(95.5) | <0.001 |
| **Race and Ethnicity (n = 70332)** |  |  |  | <0.001 |
| *White non-Hispanic* | 31291 (44.5) | 30080 (44.6) | 1211 (42.0) |  |
| *White Hispanic* | 2669 (3.8) | 2550 (3.8) | 119 (4.1) |  |
| *non-White non-Hispanic* | 30916 (44.0) | 29670 (44.0) | 1246 (43.2) |  |
| *non-White Hispanic* | 5456 (7.8) | 5146 (7.6) | 310 (10.7) |  |
| **Insurance (n = 70931)** |  |  |  | <0.001 |
| *Public* | 27711 (39.1) | 26447 (38.9) | 1264 (43.7) |  |
| *Private* | 43220 (60.9) | 41592 (61.1) | 1628 (56.3) |  |
| **$\geq 2$ Insurance Changes** | 11000 (15.3) | 10243 (14.9) | 757 (26.0) |  |
| **National ADI (n = 71677)** | 42.66 (19.52) | 42.44 (19.6) | 48.09 (17.6) | <0.001 |
| **ADI Quartile mean (SD)** |  |  |  |  |
| *Q1 (3.79 to 25.45)* | 18556 (25.9) | 18168 (26.4) | 388 (13.3) |  |
| *Q2 (25.55 to 42.76)* | 17430 (24.3) | 16833 (24.5) | 597 (20.5) |  |
| *Q3 (42.80 to 56.11)* | 17788 (24.8) | 16768 (24.4) | 1020 (35.0) |  |
| *Q4 (56.22 to 93.00)* | 17903 (25.0) | 16997 (24.7) | 906 (31.1) |  |
| **COI Group** |  |  |  | <0.001 |
| *Low/Very Low* | 18394 (25.7) | 17420 (25.3) | 974 (33.5) |  |
| *Moderate* | 13706 (19.1) | 12958 (18.8) | 748 (25.7) |  |
| *High/Very High* | 39583 (55.2) | 38394 (55.8) | 1189 (40.9) |  |
| **COI Education** |  |  |  | <0.001 |
| *Low/Very Low* | 20925 (29.2) | 19926 (29.0) | 999 (34.3) |  |
| *Moderate* | 15199 (21.2) | 14389 (20.9) | 810 (27.8) |  |
| *High/Very High* | 35559 (49.6) | 34457 (50.1) | 1102 (37.9) |  |
| **COI Health and Environment** |  |  |  | <0.001 |
| *Low/Very Low* | 3144 (4.4) | 2957 (4.3) | 187 (6.4) |  |
| *Moderate* | 12073 (16.8) | 11399 (16.6) | 674 (23.2) |  |
| *High/Very High* | 56466 (78.8) | 54416 (79.1) | 2050 (70.1) |  |
| **COI Socioeconomic** |  |  |  | <0.001 |
| *Low/Very Low* | 20845 (29.1) | 19691 (28.6) | 1154 (39.6) |  |
| *Moderate* | 11803 (16.5) | 11217 (16.3) | 586 (20.1) |  |
| *High/Very High* | 39035 (54.5) | 37864 (55.1) | 1171(40.2) |  |
| **Clinical Parameters** |  |  |  |  |
| **Premature Birth (n = 35543)** | 4550 (12.8) | 4259 (12.5) | 291 (18.5) | <0.001 |
| **BMI (n = 71457)** |  |  |  | <0.001 |
| *Underweight* | 2687 (3.8) | 2649 (3.9) | 38 (1.3) |  |
| *Healthy* | 46561 (65.1) | 45594 (66.5) | 967 (33.3) |  |
| *Overweight* | 10703 (15.0) | 10180 (14.9) | 523 (18.0) |  |
| *Obese* | 11506 (16.1) | 10130 (14.8) | 1376 (47.4) |  |
| **HTN Diagnosis** | 848 (1.2) | 177 (0.3) | 671 (23.1) | <0.001 |

Values are presented as n (%) unless otherwise indicated. National ADI is presented as mean (SD).
Percentages are column percentages. Abbreviations: n: Total participants with available data analyzed
(Parameters with no n shown had no missing data); ADI: Area Deprivation Index; COI: Child
Opportunity Index; BMI: body mass index; BP: blood pressure; Abnormal: children with at least three
abnormal BP readings on three consecutive separate visits; HTN Diagnosis: Participants with a primary
HTN diagnostic code.

The final study cohort (n = 71,683) was demographically diverse, with 44.0% identifying as non-White non-Hispanic and 11.3% as Hispanic. Nearly half were male (46.3%), and 44.0% were aged ≥13 years. Most participants were English proficient (93.6%) and had ≥2 caregivers (93.9%). The majority had private insurance (60.9%), and 15.3% experienced >2 insurance changes during the study period.

From a neighborhood perspective, 25.7% of the cohort resided in low or very low COI areas. Distribution across ADI quartiles was relatively even, with approximately one-quarter of participants in each quartile. Clinically, most children were born at term, and 31.1% had elevated BMI, including 15.0% overweight and 16.1% obese (Table 1).

### Comparison of Normal and Abnormal Blood Pressure

Participants with abnormal BP differed from those with normal BP across key demographic, clinical, and neighborhood characteristics (Table 1). Notably, a substantially higher proportion of adolescents (90.7% versus 42.0%) and children with obesity (47.4% versus 14.8%) were in the abnormal BP group compared to normal BP cohort. In addition, children with abnormal BP were more likely to experience insurance instability (26.0% versus 14.9%) and to reside in more socioeconomically disadvantaged neighborhoods, with higher mean ADI and greater representation in higher ADI quartiles. Similarly, lower neighborhood opportunity based on COI was more common among children with abnormal BP, particularly within the socioeconomic domain (39.6% versus 28.6% in low/very low COI).

### Multivariable Analysis: Abnormal Blood Pressure

In multivariable logistic regression (Table 2), the model demonstrated strong discrimination (C-statistic = 0.88). Male sex (OR 2.44, 95% CI 2.18–2.74) and adolescent age ≥13 years (OR 17.29, 95% CI 14.73–20.41) were strongly associated with abnormal BP.

**Table 2.**
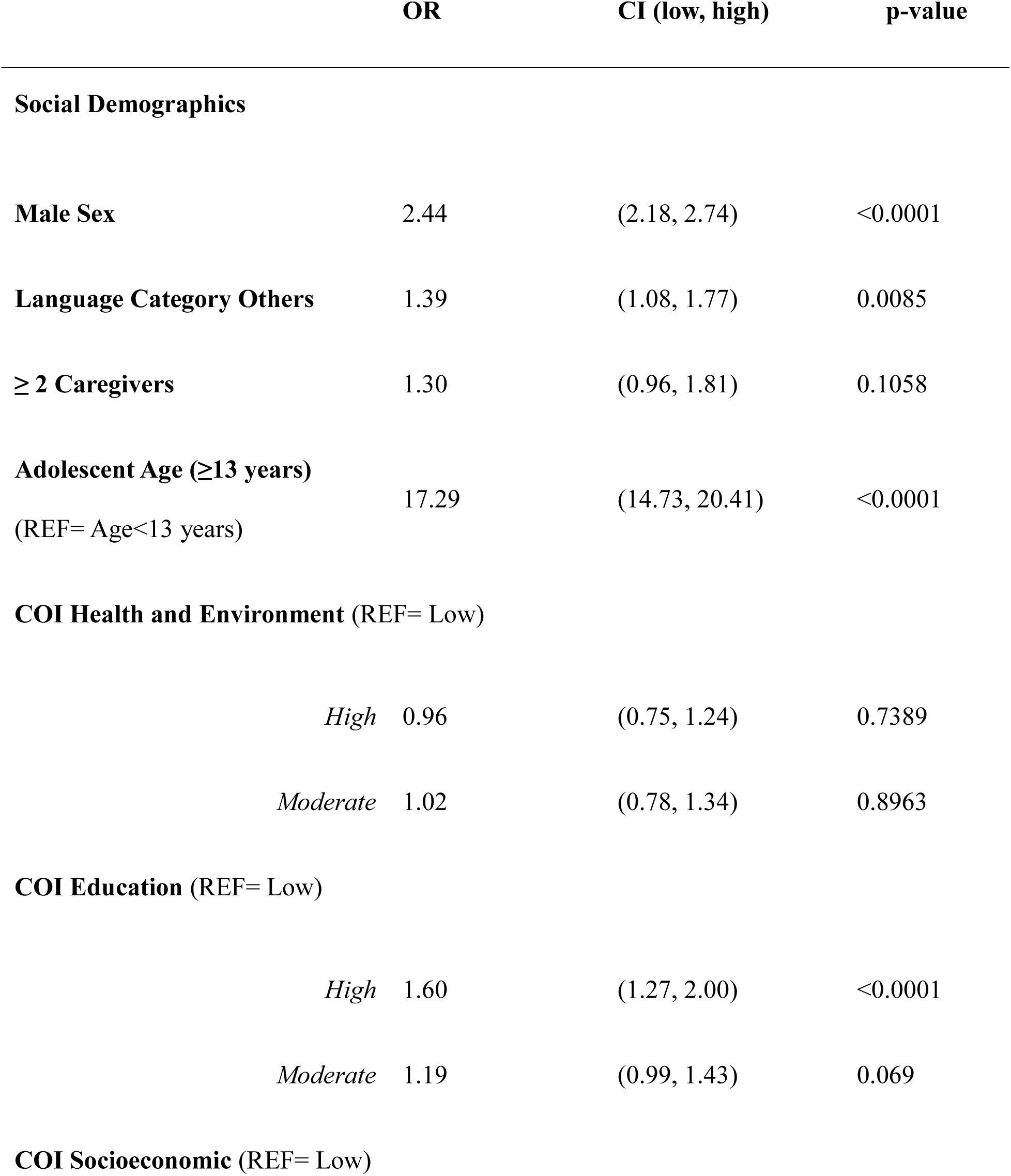
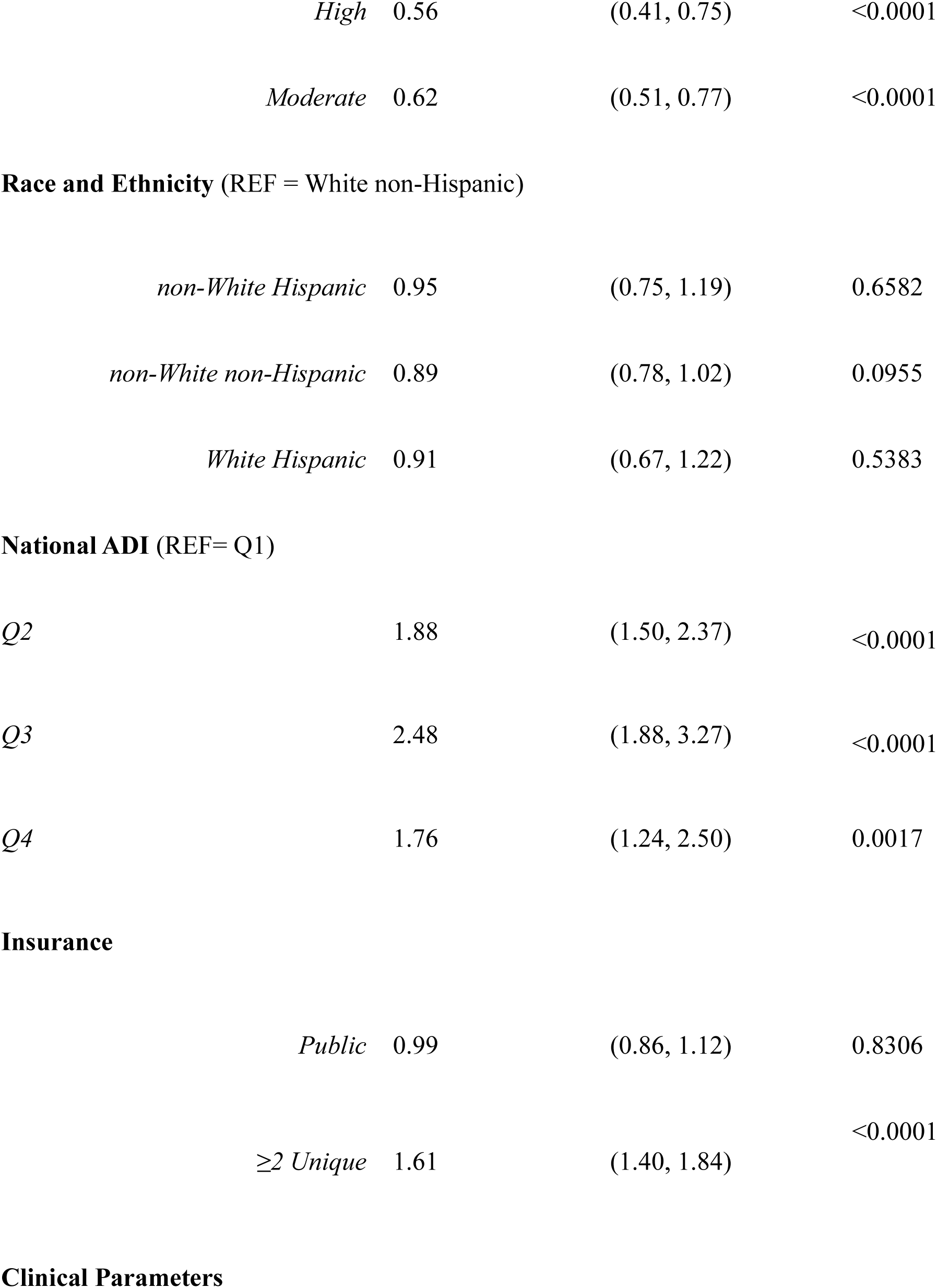
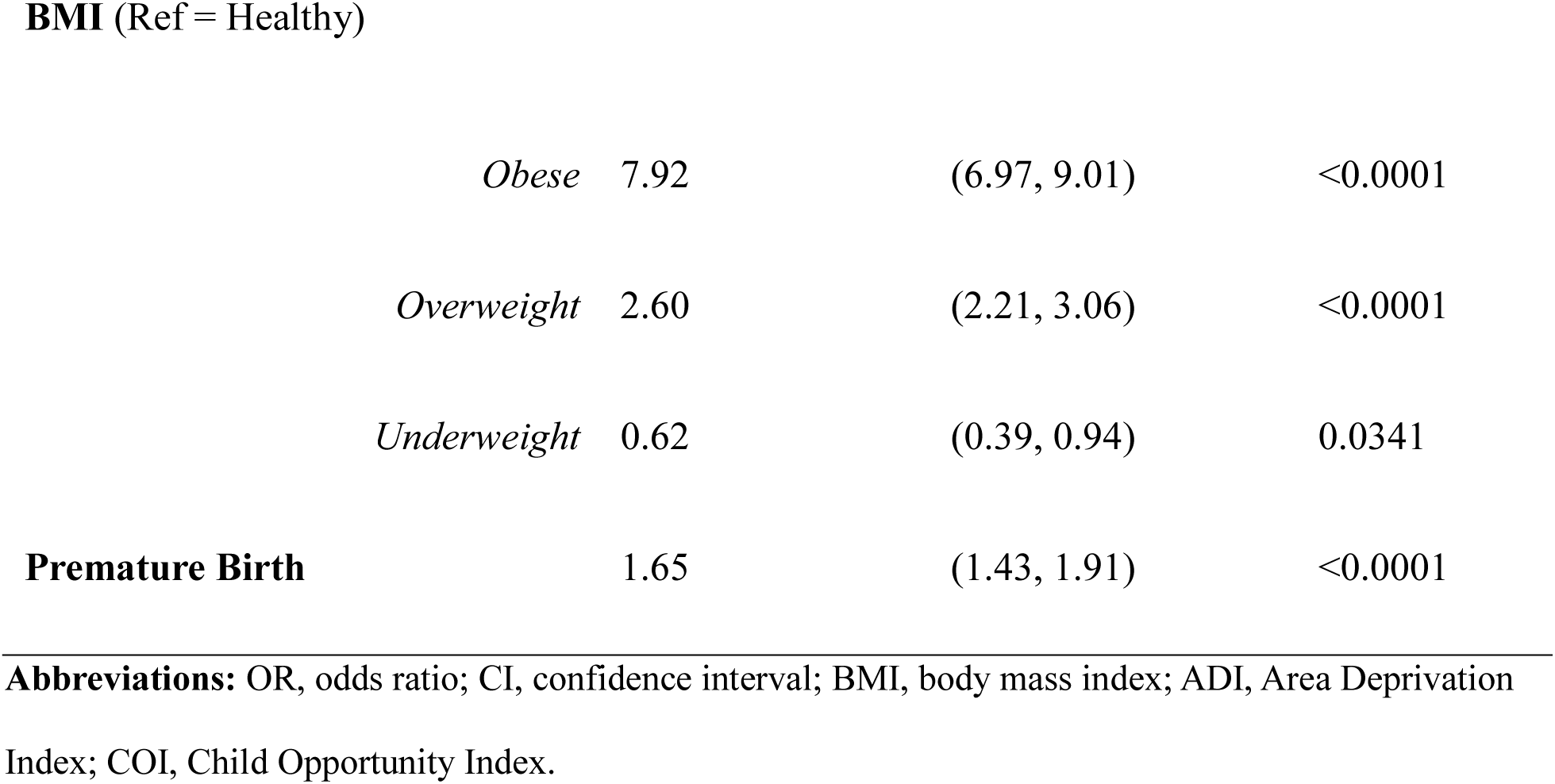
Multivariable Logistic Regression Analysis of Factors Associated with Abnormal Blood Pressure (vs Normal Blood Pressure). C-statistic- 0.88.

|  | OR | CI (low, high) | p-value |
| --- | --- | --- | --- |
| **Social Demographics** |  |  |  |
| **Male Sex** | 2.44 | (2.18, 2.74) | <0.0001 |
| **Language Category Others** | 1.39 | (1.08, 1.77) | 0.0085 |
| **≥ 2 Caregivers** | 1.30 | (0.96, 1.81) | 0.1058 |
| **Adolescent Age (≥13 years)**<br>(REF= Age<13 years) | 17.29 | (14.73, 20.41) | <0.0001 |
| **COI Health and Environment (REF= Low)** |  |  |  |
| *High* | 0.96 | (0.75, 1.24) | 0.7389 |
| *Moderate* | 1.02 | (0.78, 1.34) | 0.8963 |
| **COI Education (REF= Low)** |  |  |  |
| *High* | 1.60 | (1.27, 2.00) | <0.0001 |
| *Moderate* | 1.19 | (0.99, 1.43) | 0.069 |
| **COI Socioeconomic (REF= Low)** |  |  |  |
| *High* | 0.56 | (0.41, 0.75) | <0.0001 |
| *Moderate* | 0.62 | (0.51, 0.77) | <0.0001 |
| **Race and Ethnicity (REF = White non-Hispanic)** |  |  |  |
| *non-White Hispanic* | 0.95 | (0.75, 1.19) | 0.6582 |
| *non-White non-Hispanic* | 0.89 | (0.78, 1.02) | 0.0955 |
| *White Hispanic* | 0.91 | (0.67, 1.22) | 0.5383 |
| **National ADI (REF= Q1)** |  |  |  |
| *Q2* | 1.88 | (1.50, 2.37) | <0.0001 |
| *Q3* | 2.48 | (1.88, 3.27) | <0.0001 |
| *Q4* | 1.76 | (1.24, 2.50) | 0.0017 |
| **Insurance** |  |  |  |
| *Public* | 0.99 | (0.86, 1.12) | 0.8306 |
| *≥2 Unique* | 1.61 | (1.40, 1.84) | <0.0001 |
| **BMI (Ref = Healthy)** |  |  |  |
| *Obese* | 7.92 | (6.97, 9.01) | <0.0001 |
| *Overweight* | 2.60 | (2.21, 3.06) | <0.0001 |
| *Underweight* | 0.62 | (0.39, 0.94) | 0.0341 |
| **Premature Birth** | 1.65 | (1.43, 1.91) | <0.0001 |

**Abbreviations:** OR, odds ratio; CI, confidence interval; BMI, body mass index; ADI, Area Deprivation Index; COI, Child Opportunity Index.

BMI demonstrated a strong association, with overweight (OR 2.60, 95% CI 2.21–3.06) and obesity (OR 7.92, 95% CI 6.97–9.01) significantly increasing the odds of abnormal BP, while underweight status was associated with lower odds (OR 0.62, 95% CI 0.39–0.94). Prematurity also was independently associated with abnormal BP (OR 1.65, 95% CI 1.43–1.91).

Insurance instability (≥2 insurance changes) was associated with increased odds of abnormal BP (OR 1.61, 95% CI 1.40–1.84), whereas insurance type and race/ethnicity were not significant. ADI demonstrated a graded association, with increasing odds across quartiles compared with Q1, including Q2 (OR 1.88, 95% CI 1.50–2.37), Q3 (OR 2.48, 95% CI 1.88–3.27), and Q4 (OR 1.76, 95% CI 1.24–2.50). Within COI domains, higher educational opportunity was associated with increased odds of abnormal BP (OR 1.60, 95% CI 1.27–2.00), whereas higher socioeconomic opportunity was associated with lower odds (OR 0.56, 95% CI 0.41–0.75). The health and environment domain was not significantly associated.

### Multivariable Analysis: Stage 1 and Stage 2 Hypertension

The model restricted to stage 1 or 2 HTN (Table 3) also demonstrated strong discrimination (C-statistic = 0.90). Male sex (OR 2.70, 95% CI 2.07–3.55) and adolescent age ≥13 years (OR 14.69, 95% CI 10.35–21.49) were strongly associated with stage 1/2 HTN.

**Table 3.**
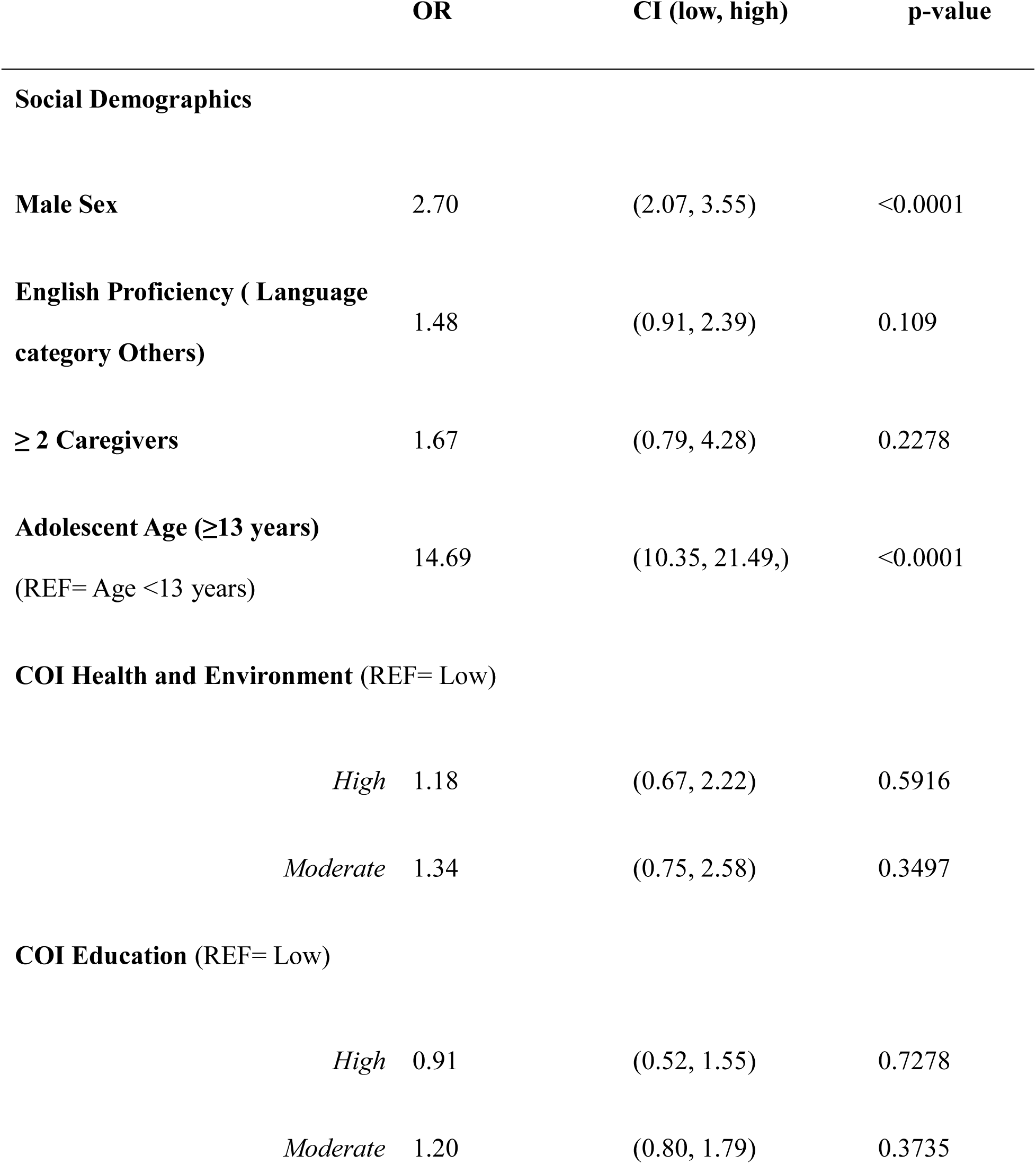
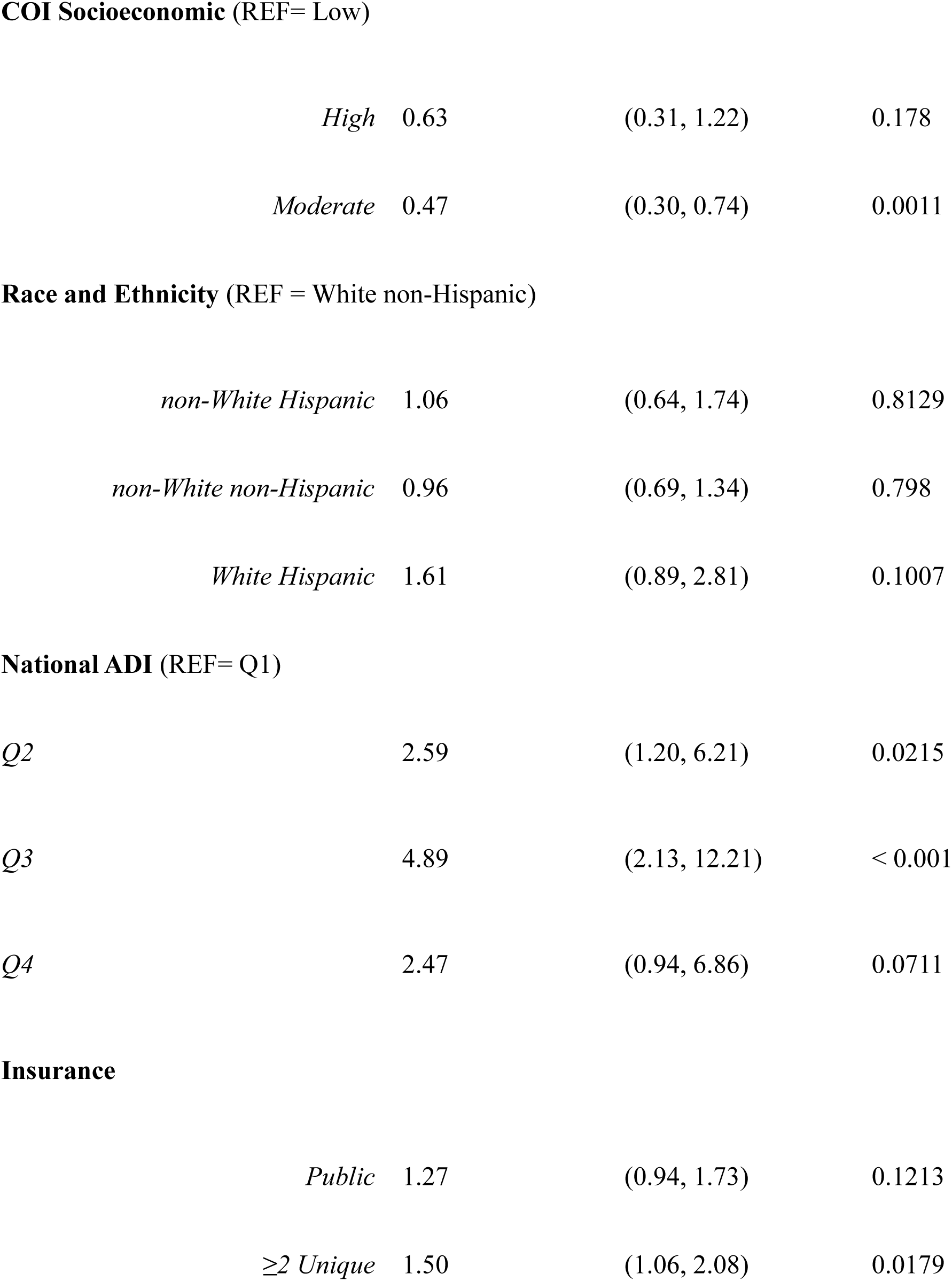
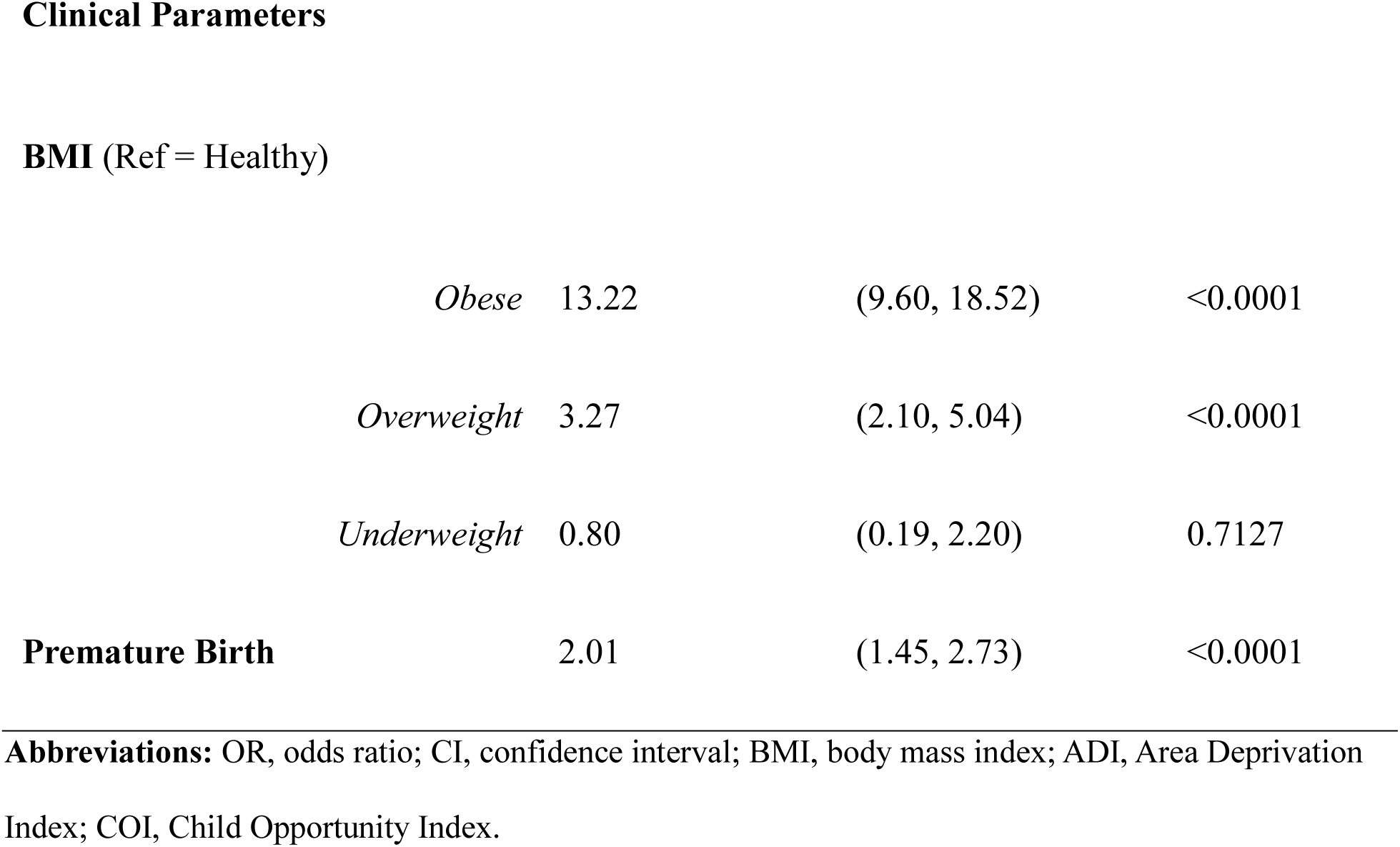
Multivariable Logistic Regression Analysis of Factors Associated with Stage 1/Stage 2 Hypertension (vs Normal Blood Pressure). C-statistic- 0.90.

|  | OR | CI (low, high) | p-value |
| --- | --- | --- | --- |
| **Social Demographics** |  |  |  |
| **Male Sex** | 2.70 | (2.07, 3.55) | <0.0001 |
| **English Proficiency ( Language category Others)** | 1.48 | (0.91, 2.39) | 0.109 |
| **≥ 2 Caregivers** | 1.67 | (0.79, 4.28) | 0.2278 |
| **Adolescent Age (≥13 years)**<br>(REF= Age <13 years) | 14.69 | (10.35, 21.49,) | <0.0001 |
| **COI Health and Environment (REF= Low)** |  |  |  |
| *High* | 1.18 | (0.67, 2.22) | 0.5916 |
| *Moderate* | 1.34 | (0.75, 2.58) | 0.3497 |
| **COI Education (REF= Low)** |  |  |  |
| *High* | 0.91 | (0.52, 1.55) | 0.7278 |
| *Moderate* | 1.20 | (0.80, 1.79) | 0.3735 |
| **COI Socioeconomic** (REF= Low) |  |  |  |
| *High* | 0.63 | (0.31, 1.22) | 0.178 |
| *Moderate* | 0.47 | (0.30, 0.74) | 0.0011 |
| **Race and Ethnicity** (REF = White non-Hispanic) |  |  |  |
| *non-White Hispanic* | 1.06 | (0.64, 1.74) | 0.8129 |
| *non-White non-Hispanic* | 0.96 | (0.69, 1.34) | 0.798 |
| *White Hispanic* | 1.61 | (0.89, 2.81) | 0.1007 |
| **National ADI** (REF= Q1) |  |  |  |
| *Q2* | 2.59 | (1.20, 6.21) | 0.0215 |
| *Q3* | 4.89 | (2.13, 12.21) | < 0.001 |
| *Q4* | 2.47 | (0.94, 6.86) | 0.0711 |
| **Insurance** |  |  |  |
| *Public* | 1.27 | (0.94, 1.73) | 0.1213 |
| *≥2 Unique* | 1.50 | (1.06, 2.08) | 0.0179 |
| BMI (Ref = Healthy) |  |  |  |
| *Obese* | 13.22 | (9.60, 18.52) | <0.0001 |
| *Overweight* | 3.27 | (2.10, 5.04) | <0.0001 |
| *Underweight* | 0.80 | (0.19, 2.20) | 0.7127 |
| **Premature Birth** | 2.01 | (1.45, 2.73) | <0.0001 |

**Abbreviations:** OR, odds ratio; CI, confidence interval; BMI, body mass index; ADI, Area Deprivation Index; COI, Child Opportunity Index.

BMI remained the strongest clinical predictor, with obesity (OR 13.22, 95% CI 9.60–18.52) and overweight status (OR 3.27, 95% CI 2.10–5.04) significantly increasing the odds of stage 1/2 HTN. Prematurity also was independently associated (OR 2.01, 95% CI 1.45–2.73).

Insurance instability was independently associated with stage 1/2 HTN (OR 1.50, 95% CI 1.06–2.08), while language, number of caregivers reported, race/ethnicity, and insurance type were not significantly associated. ADI remained associated with stage 1/2 HTN, particularly for Q2 (OR 2.59, 95% CI 1.20–6.21) and Q3 (OR 4.89, 95% CI 2.13–12.21), whereas Q4 did not reach statistical significance. In contrast to the abnormal BP model, most COI domains were not significantly associated, except for moderate socioeconomic opportunity, which was associated with lower odds of stage 1/2 HTN (OR 0.47, 95% CI 0.30–0.74).

## DISCUSSION

In this large cohort of children with longitudinal continuity of primary care, 4% met physiological criteria for HTN based on repeated BP measurements, consistent with the reported prevalence of HTN, yet lacked a documented diagnosis, highlighting persistent under recognition of HTN in pediatric primary care practice.^1,2,11,33–35^ Across both abnormal BP and stage 1 and stage 2 HTN models, adolescent age, male sex, obesity, prematurity, and insurance instability were strongly associated with persistent BP elevation. Importantly, structural factors, including neighborhood deprivation and opportunity, were associated with BP outcomes even for patients who maintained continuity of care, indicating that access alone does not eliminate disparities in risk or recognition.

The associations between obesity, male sex, and adolescent age with abnormal BP and HTN are consistent with prior pediatric literature demonstrating the central role of adiposity, sex, and pubertal physiology in BP regulation.^1,2,6,7,18,36^ In particular, elevated BMI demonstrated an incremental relationship, with increasing weight conferring markedly increased odds of both abnormal BP and stage 1/2 HTN.^2,18,36^ These findings reinforce well-established cardiometabolic pathways linking adiposity to vascular dysfunction, sympathetic activation, and early end-organ effects.^5,6,13,14,16,37–42^ Similarly, the lack of a consistent independent association between race/ethnicity and BP outcomes after adjustment aligns with emerging evidence suggesting that previously observed disparities may be mediated by structural and socioeconomic factors rather than race itself, when more comprehensive measures of socioeconomic disparities and opportunities are included.^21,22,36^ In addition, neighborhood deprivation and opportunity are influenced by historical and contemporary structural factors, including residential segregation and structural racism, which may contribute to differences in cardiovascular risk and healthcare experiences independent of individual characteristics.

Our findings extend prior work by leveraging longitudinal BP data and characterizing BPs according to established guidelines to determine possible HTN diagnosis, rather than relying on diagnostic codes or single-visit BP measurements.^1,2,24,43^ Even among children who maintained continuity of care, higher ADI was associated with increased odds of abnormal BP and stage 1/2 HTN, suggesting that structural disadvantage may influence cardiometabolic risk independent of access to pediatric primary care.^21^ This suggests that ongoing contact with the healthcare system was insufficient to mitigate underlying risk or ensure timely recognition.

COI provides additional insight into factors that may influence the risk of abnormal BP. Higher opportunity within the education domain was associated with increased odds of abnormal BP. Although the mechanism is unclear, this finding may reflect differential healthcare engagement and surveillance rather than true disease burden, whereby children in higher-opportunity neighborhoods have more opportunities for repeat BP assessment and identification of persistent abnormalities. In contrast, higher socioeconomic opportunity was associated with lower odds of abnormal BP, consistent with reduced underlying cardiometabolic risk, access to healthy foods, and likely less stress in more resource-rich environments^27,44^; however, these associations were attenuated when analyses were restricted to children with stage 1/2 HTN, where underlying biological and clinical factors appear to play a more dominant role.

Insurance instability further contextualizes these findings.^20,45–47^ Children with multiple insurance changes had higher odds of abnormal BP and stage 1/2 HTN, suggesting exposure to ongoing structural and financial instability. At the same time, their continuity in care indicates an ability, or necessity, to navigate the healthcare system despite these disruptions. Considered together, these patterns point to a more complex dynamic in which structural vulnerability and healthcare engagement coexist, shaping both the possible development of HTN and the likelihood that it is detected.

These findings have important implications for clinical practice and health system design. First, they demonstrate that current approaches to pediatric HTN detection, largely dependent on clinic-based BP measurements, clinician interpretation, and follow up, are insufficient for HTN detection, even among patients who maintain continuity of care.^46–48^ This suggests that the population captured in this study represents only a subset of affected children, as those who do not return for follow up are even less likely to be identified. Second, association between insurance instability and persistent abnormal BP highlights continuity of coverage as a key, potentially modifiable systems-level factor influencing both longitudinal care and HTN detection.

Addressing these gaps will require moving beyond one-size-fits-all strategies toward more tailored, systems-based approaches to HTN recognition. Such approaches should integrate clinical risk factors (e.g., obesity, prematurity, adolescence) with social context, including neighborhood deprivation and patterns of healthcare engagement. For children who remain in care yet continue to have unrecognized HTN, health systems should implement automated EMR-based tools to identify repeated BP elevations, establish standardized clinical pathways for evaluation and follow up, and incorporate multidisciplinary resources such as nutrition, social work, and community-based support, to mitigate social factors implicated in HTN risk and detection.^46^ At the same time, strategies to improve access remain essential. Children experiencing insurance instability or residing in lower-opportunity environments may face barriers to consistent care, and efforts to expand access through out of office BP monitoring including school-based screening, community partnerships, home BP monitoring, or remote BP monitoring, are critical to ensuring at-risk populations are reached.

### Limitations

Several limitations should be considered. First, this was a retrospective study using EMR data, which may be subject to misclassification and incomplete capture of clinical and social variables. Although we used repeated BP measurements across visits to approximate guideline-based diagnostic criteria, BP was obtained in routine clinical settings and may be influenced by measurement variability, technique, or situational factors such as anxiety. Second, our cohort was restricted to children with at least three primary care visits, which was necessary to assess longitudinal BP patterns but limits generalizability to children with limited or fragmented care, who may be at even greater risk for under recognition. Third, neighborhood-level measures such as ADI and COI were assigned based on zip code and may not fully capture individual-level socioeconomic conditions. In addition, COI domains are interrelated and may reflect overlapping pathways, limiting causal interpretation. Fourth, although we examined HTN diagnostic codes to assess detection, coding practices may vary across providers and may not fully reflect clinical decision-making. Finally, the electronic medical record contained limited race and ethnicity classifications, requiring aggregation into broader categories for analysis. This approach may have masked important differences among individual racial and ethnic groups and limited our ability to identify disparities within these populations.

## Conclusions

In this large multistate cohort of children with longitudinal continuity of primary care engagement, a substantial proportion met physiological criteria for HTN based on repeated BP measurements yet lacked a documented diagnosis, highlighting persistent under recognition even among patients with continuity of care. Clinical factors such as obesity, age, and prematurity are strong drivers of risk for abnormal BP; however, social and structural determinants, including neighborhood deprivation, opportunity, and insurance instability, continue to shape the risk and recognition of HTN.

These findings demonstrate that coordinated, system-level approaches are needed that enhance recognition within clinical workflows while expanding access to care across community settings. Integrating longitudinal BP tracking, standardized clinical pathways, and multidisciplinary support with strategies such as school-based screening, home BP monitoring, and community partnerships may improve early identification and management. Addressing pediatric HTN will require a multi-pronged approach that simultaneously accounts for clinical risk, social context, and healthcare system change to ensure timely and equitable detection.

### Perspectives

This study highlights that under recognition of pediatric hypertension persists even among children who maintain continuity of care, demonstrating limitations in current clinic-based detection strategies. Our findings suggest social factors continue to influence hypertension risk and its recognition, even when access to care is present. This shifts the focus from access alone toward the need for improved operationalization of longitudinal blood pressure assessment within health systems. Future efforts should prioritize automated BP tracking integration, standardized clinical pathways, and population health tools that identify at-risk patients in real time. At the same time, expanding access through community-based approaches, including school-based screening, home BP monitoring, and remote care, will be essential to reach children with fragmented or absent care. Together, these findings support a broader systems-level approach that aligns clinical workflows with social context to improve early identification and management of pediatric hypertension.

## NOVELTY AND RELEVANCE

### What Is New?

- Uses longitudinal BP measurements across multiple visits to identify hypertension rather than relying on diagnostic codes
- Focuses on children with continuity of care to isolate effects beyond healthcare access
- Demonstrates that neighborhood factors (ADI, COI) and insurance instability remain associated with hypertension even among engaged patients

### What Is Relevant?

- Highlights persistent gaps in pediatric hypertension recognition despite ongoing healthcare access
- Identifies social and health system structural factors that influence hypertension risk and detection
- Emphasizes limitations of current clinic-based screening approaches

### Clinical/Pathophysiological Implications

- Supports the need for system-level strategies to improve hypertension detection within clinical workflows
- Reinforces the importance of integrating clinical care with community-based approaches to improve equitable hypertension identification

## Data Availability

The data, analytic methods, and study materials that support the findings of this study are available from the corresponding author upon reasonable request, subject to institutional and IRB requirements to protect participant confidentiality

## Acknowledgments

The authors acknowledge the assistance of Editorial Services of Nemours Children’s Health.

## Sources of funding

None

## Disclosures

None

## Non-standard Abbreviations and Acronyms

ADI: Area Deprivation Index
COI: Child Opportunity Index
EMR: Electronic Medical Record
HTN: Hypertension
SDOH: Social Drivers of Health

## Notes

### Competing Interest Statement

The authors have declared no competing interest.

### Funding Statement

No external funding was received for this work. The authors and their institutions did not receive payment or services from a third party for any aspect of the submitted work, including study design, data collection, data analysis, data interpretation, or manuscript preparation.

### Author Declarations

The study was reviewed and approved by the Nemours Children's Health Institutional Review Board. The requirement for informed consent was waived because the study involved retrospective analysis of existing electronic medical record data using de-identified data.

